# The Net Clinical Benefit of Fondaparinux vs LMWH in Chinese Non-ST-Segment Elevation Acute Coronary Syndrome Patients: the REFOCAS Nonrandomized Controlled Trial

**DOI:** 10.1101/2025.02.24.25322821

**Authors:** Dandan Li, Xiaodan Tuo, Yani Yu, Genshan Ma, Chuanyu Gao, Wei Mao, Peng Qu, Limin Liu, Zhuo Shang, Xunli Yin, Guozhen Wang, Aifen Liu, Yundai Chen

## Abstract

**Background and aims:** In Chinese patients with non-ST-segment elevation acute coronary syndrome (NSTE-ACS), it is essential to evaluate the net clinical benefit (NCB) of fondaparinux compared to low molecular weight heparin (LMWH) in NSTE-ACS patients with varying bleeding and ischemic risks.

**Methods:** This multicenter, prospective, open-label, real-world study enrolled 8066 adult patients undergoing non-emergency percutaneous coronary intervention (PCI) at 88 hospitals from July 2019 to July 2021. Patients received fondaparinux (2.5 mg/d) or LMWH (1 mg/kg, twice/day) were followed up within 6 months. The GRACE and CRUSADE scores were used to stratify the included patients. The primary outcome was the incidence of NCB event (all-cause death, reinfarction, nonfatal stroke, or BARC≥ type 2 bleeding) at 30 days.

**Results:** A total of 5430 patients received fondaparinux and 2636 received LMWH. The NCB outcome occurred in 136 patients (2.5%) receiving fondaparinux and in 110 patients (4.2%) receiving LMWH [HR=0.594 (95% CI: 0.462, 0.764), p<0.001]. The 30-day incidence of BARC≥2 bleeding was significantly lower in the fondaparinux group compared with the LMWH group [68 (1.3%) vs. 66 (2.5%), HR=0.498 (95% CI: 0.355, 0.699), p<0.001].Among patients with lower risk, defined by GRACE score≤140 and CRUSADE scores≤40, fondaparinux significantly reduced the incidence of MACE and BARC ≥ 2 bleeding. Fondaparinux decreased the risk of BARC≥2 bleeding across different risk groups, with absolute risk reductions (ARD) of -0.9% for CRUSADE score≤40, -3.0% for CRUSADE score>40, -1.2% for GRACE score≤140, and -1.7% for GRACE score>140, indicating a more pronounced protective effect in individuals at high bleeding risk.

**Conclusions:** In Chinese patients with NSTE-ACS, fondaparinux effectively diminished the incidence of the NCB event of 30 days compared with LMWH. Additionally, fondaparinux exhibited superior safety than LMWH in patients with high bleeding risk, consistently outperformed LMWH in terms of efficacy and safety in the low-risk group as well.

## Introduction

Patients diagnosed with Non-ST-segment elevation acute coronary syndromes (NSTE-ACS) should receive parenteral anticoagulation, based on the assessment of ischemia and bleeding risks, particularly during revascularization therapy[1–3]. In China, the currently available perioperative parenteral anticoagulants include unfractionated heparin (UFH), low-molecular-weight heparin (LMWH), fondaparinux, and bivalirudin[4,5], among others. However, it is currently uncertain whether the clinical application of anticoagulants aligns with established guidelines and the current status of anticoagulant use in the perioperative period of percutaneous coronary intervention (PCI).

Fondaparinux is the first synthetic selective inhibitor of factor Xa, used in the treatment of various thrombotic disorders[6]. Previous Large randomized controlled trials (RCTs) and real-world studies[7,8] have previously demonstrated that fondaparinux significantly reduced the risk of in-hospital major bleeding among NSTE-ACS patients compared to enoxaparin. This reduction was associated with decreased short-term and long-term mortality rates. However, these studies [7,8] were completed before 2010, and the populations enrolled and treatment regimens used differed from those in current cardiovascular disease management. Notable differences include variations in cardiovascular risk factors, patient age demographics, and the use of dual antiplatelet and medication escalation strategies[9]. Most previous studies were conducted in European and American populations, whereas East Asian populations have different risk profiles for ischemia or bleeding following PCI[10,11], typically exhibiting lower ischemia risk and higher bleeding risk.

The Net Clinical Benefit (NCB) can be utilized for assessing the net benefit of different treatment in terms of ischemic and bleeding risks, thereby directing the application of anticoagulants and potent antiplatelet drugs. Moreover, the GRACE and CRUSADE scores for ischemia and bleeding risk were used to stratify ACS patients[12,13]. However, there has been limited discussion about the use of anticoagulation drugs in patients with varying risk stratification. Therefore, we stratified patient cohort using GRACE and CRUSADE scores and evaluated the incidence of ischemic and bleeding events in Chinese NSTE-ACS patients undergoing non-emergency PCI therapy with various antithrombotic regimens: fondaparinux or LMWH combined with ticagrelor or clopidogrel. We aimed to evaluate the comparative effects of fondaparinux versus LMWH on major bleeding and cardiovascular events in NSTE-ACS patients, to inform anticoagulant therapy decisions in NSTE-ACS patients with varying bleeding and ischemic risks.

## Methods

### Study design and participants

The REFOCAS study is a multicenter, prospective, open-label, real-world clinical trial conducted in China, and screened 8209 patients from 88 hospitals nationwide from July 2019 to July 2021 for eligibility (registration number: ChiCTR1900025366; www.chictr.org.cn). Following the provision of informed consent, all eligible patients diagnosed with NSTE-ACS were enrolled at a 2:1 allocation ratio into the fondaparinux group or LMWH group at each participating center, without blinding of patients or investigators. Patients were followed up for six months.

The study was approved by the Ethics Committee of the General Hospital of the Chinese People’s Liberation Army (approval number S201915001) and was conducted according to the principles of the Declaration of Helsinki. All patients signed an informed consent form before enrollment. Inclusion criteria were as follows: (1) age >18 years; (2) NSTE-ACS patients undergoing non-emergency PCI treatment, including those with Non-ST-segment elevation myocardial infarction (NSTEMI) and UA; (3) subjects or their legal representatives were informed about the study’s nature and protocol provisions, ensured their compliance, and signed the informed consent form. Exclusion criteria included: (1) patients with ST-segment elevation myocardial infarction; (2) patients with NSTE-ACS undergoing emergent PCI therapy; (3) patients with allergy or contraindications to fondaparinux and LMWH; (4) patients had already participated in another drug/device study or were enrolled in another drug/device study during the follow-up period; (5) patients deemed potentially non-compliant with the study protocol, including the follow-up period, due to conditions such as psychiatric abnormalities, alcohol or drug abuse. Patients were excluded if they were enrolled and used fondaparinux and LMWH simultaneously, or if they did not use any antiplatelet drugs.

### Procedures

The LMWH group received LMWH a dose of 1 mg/kg twice daily, with no restrictions on the choice of manufacturer. For patients with NSTE-ACS already treated with LMWH, no additional dose was needed if the last subcutaneous injection was administered less than 8 hours before PCI. Otherwise, an additional LMWH dose of 0.3 mg/kg was required be administered intravenously. Switching to other types of anticoagulants at the time of PCI is not recommended[2]. For patients with severe renal insufficiency [creatinine clearance (Ccr) <30 ml/min], the LMWH dosing schedule needed to be adjusted to once daily. For patients with moderate and mild renal insufficiency, close monitoring during treatment was recommended. Ccr can be calculated using the following formula: (140 - age) ×weight (kg)/72 × blood creatinine (mg/dl). For females, multiply the result by 0.85.

Fondaparinux (2.5 mg daily, Jiangsu Hengrui Pharmaceutical Co., Ltd) was administered subcutaneously for a duration of 2-8 days or until hospital discharge. For patients undergoing PCI with fondaparinux, a single intraoperative intravenous push of UFH 85 IU/kg or 60 IU/kg in combination with GP IIb/IIIa receptor inhibitors, was recommended[14]. Notably, no dose adjustment was necessary for patients with acute or chronic kidney disease. Fondaparinux was not recommended for use in patients with a Ccr less than 20 ml/min.

Upon admission, we assessed all patients’ GRACE and CRUSADE scores[12,13], defining the high-risk group as those with a GRACE score greater than 140 and a CRUSADE greater than 40. In accordance with current guidelines and considering individual patient characteristics, all patients received appropriate dual antiplatelet therapy, which included aspirin combined with either clopidogrel or ticagrelor. The clinician determined the duration of in-hospital anticoagulation, the surgical puncture site, and the type of stent to be used. The patients underwent telephone follow-up assessments 30 days and 6 months after enrollment.

### Outcomes

The primary study endpoint was the incidence of NCB event[15] (all-cause death, reinfarction, nonfatal stroke, or BARC ≥2 bleeding) at 30 days of enrollment. Secondary endpoints included Major Adverse Cardiovascular Events (MACE) at hospitalization, 30 days, and six months, which comprised all-cause death, reinfarction, and nonfatal stroke. Endpoint events and BARC staging definitions are shown in the Appendix. The Clinical Events Committee (CEC), established for this study, was composed of independent, non-participating experts in cardiac intervention. The CEC was responsible for reviewing and classifying all reported events.

### Statistical analysis

Based on existing literature, it was assumed that the incidence of 30-day composite endpoint events in both groups would be 14%. The significance threshold was set at 0.025 (one-sided), coupled with a non-inferiority margin of 2.8%, ensuring a test power of 90%. The estimated sample size was 4842 cases in the Fondaparinux group and 2421 cases in the LMWH group, totaling 7263 patients. In practice, 7453 patients were ultimately enrolled and completed the trial. Statistical analysis was conducted using SPSS 26.0 and R software, with descriptive statistics performed for all data. Quantitative data are expressed as mean ± standard deviation or median with

interquartile range and analyzed using t-tests. Categorical data are presented as percentages and analyzed using the chi-square or Fisher’s exact tests. A two-sided test was chosen, with 0.05 as the test level and *P*<0.05 considered statistically significant. Baseline characteristics were described and compared between groups. Primary and secondary outcomes were described in groups and compared between groups using the chi-square test or the exact probability method. Survival analysis was performed using the Kaplan-Meier method to calculate survival rates. Cox proportional hazard models were used to calculate hazard ratios (HRs) and two-sided 95% confidence intervals (CIs). Additionally, we conducted analyses to assess patient prognosis during hospitalization, at 30 days, and at 180 days post-procedure. We calculated the absolute risk differences within the population and performed subgroup analyses based on predetermined cardiovascular risk factors, including age, gender, BMI, Ccr, heart function classification, medical history (hypertension, diabetes, prior PCI), postoperative anticoagulation use, and in-hospital dual antiplatelet therapy. Safety and efficacy analyses were carried out on the intention-to-treat population, and sensitivity analyses were performed on the per-protocol population. The detailed calculations are described in Appendix 2 of the Statistical analysis plan.

## Results

### Baseline characteristics

A total of 5430 patients were treated with fondaparinux and 2636 with LMWH, with 4976 and 2477, respectively, completing the trial. The characteristics of the patients and their therapies are listed in Table 1. Patients treated with fondaparinux and LMWH had the same median age of 66 years. The fondaparinux group exhibited a lower prevalence of previous PCIs at 20.8% versus 21.8% in the LMWH group, and a higher proportion of patients with a prior diagnosis of myocardial infarction (13.6% vs. 12.7%). The GRACE score, which indicates the proportion of high-ischemic-risk patients, was consistent between the two groups at 20.6%, whereas the fondaparinux group had a lower proportion of high-bleeding-risk patients according to the CRUSADE scores (16.2% vs. 17.2%). Regarding in-hospital therapies, a higher percentage of patients underwent PCI in the fondaparinux group (43.6%) compared to the LMWH group (42.6%). Although the baselines of the two groups were not identical, there were no significant statistical differences, indicating good comparability between the two groups.

**Table 1:** Baseline characteristics of the study groups.

|  | LMWH<br>(n=2636) | Fondaparinux<br>(n=5430) | p value |
| --- | --- | --- | --- |
| Age, years | 66.00(58.00, 73.00) | 66.00(58.00, 74.00) | 0.505 |
| <75 years | 2069 (78.5 %) | 4214 (77.6 %) | 0.375 |
| ≥75years | 567 (21.5 %) | 1216 (22.4 %) |  |
| Sex |  |  | 0.214 |
| Male | 1582 (60.0 %) | 3180 (58.6 %) |  |
| Female | 1054 (40.0 %) | 2250 (41.4 %) |  |
| Diagnosis at study entry |  |  | 0.437 |
| UA | 1826 (69.3 %) | 3715 (68.4 %) |  |
| NSTEMI | 810 (30.7 %) | 1715 (31.6 %) |  |
| BMI, kg/m<sup>2</sup> | 24.61(22.66, 26.99) | 24.68(22.58, 26.89) | 0.832 |
| ≥25 | 1188 (45.1 %) | 2429 (45.4 %) | 0.794 |
| < 25 | 1448 (54.9 %) | 2904 (54.6 %) |  |
| Hypertension | 1721 (65.3 %) | 3610 (66.5 %) | 0.288 |
| Diabetes | 766 (29.1 %) | 1565 (28.8 %) | 0.825 |
| Smoking | 824 (31.3 %) | 1628 (30.0 %) | 0.242 |
| Previous myocardial infarction | 336 (12.7 %) | 736 (13.6 %) | 0.316 |
| Previous percutaneous coronary intervention | 574 (21.8 %) | 1132 (20.8 %) | 0.338 |
| Previous stroke | 432 (16.4 %) | 925 (17.0 %) | 0.467 |
| Killip class* |  |  | 0.998 |
| I | 2290 (86.9 %) | 4727 (87.1 %) |  |
| II | 238 (9.0 %) | 486 (9.0 %) |  |
| III | 77 (2.9 %) | 156 (2.9 %) |  |
| IV | 28 (1.1 %) | 56 (1.0 %) |  |
| Heart rate, beats/min | 74.00(66.00, 84.00) | 74.00(66.00, 84.00) | 0.317 |
| Systolic pressure, mmHg | 138.00(125.00, 153.00) | 138.00(125.00, 153.00) | 0.667 |
| Diastolic pressure, mmHg | 80.00(73.75, 90.00) | 80.00(73.00, 90.00) | 0.618 |
| Hemoglobin, g/L | 135.00(123.00, 147.00) | 135.00(124.00, 146.00) | 0.580 |
| HCT, % | 40.30(37.00, 43.40) | 40.25(37.20, 43.40) | 0.551 |
| PLT, 10<sup>9</sup>/L | 206.00(171.25, 247.00) | 205.00(171.00, 245.00) | 0.294 |
| WBC,10<sup>9</sup>/L | 6.57(5.34, 8.00) | 6.51(5.40, 7.94) | 0.478 |
| eGFR, mL/min per 1.73 m<sup>2</sup> | 96.01(78.01, 115.00) | 96.76(78.83, 115.08) | 0.600 |
| GRACE > 140 | 547 (20.8 %) | 1130 (20.8 %) | 0.951 |
| GRACE≤140 | 2089 (79.2 %) | 4300 (79.2 %) | 0.951 |
| CRUSADE>40 | 457 (17.3 %) | 881 (16.2 %) | 0.208 |
| CRUSADE≤40 | 2179 (82.7 %) | 4549 (83.8 %) | 0.208 |
| PCI | 1124 (42.6 %) | 2366 (43.6 %) | 0.428 |
| CABG | 6 (0.2 %) | 18 (0.3 %) | 0.422 |
| Aspirin+Clopidogrel | 1433 (54.4 %) | 2926 (53.9 %) | 0.687 |
| Aspirin+Ticagrelor | 1107 (42.0 %) | 2318 (42.7 %) | 0.555 |
| Beta-blocker | 1777 (67.4 %) | 3544 (65.3 %) | 0.056 |
| Statins | 2393 (90.8 %) | 4803 (88.5 %) | 0.002 |
| ACEI/ARB | 1164 (44.2 %) | 2311 (42.6 %) | 0.174 |

Data are shown as n (%) or median (IQR). BMI=body mass index. \*Killip class was defined as follows: Class I, no clinical signs of heart failure; Class II, rales or crackles in the lungs, presence of a third heart sound, or an elevated jugular venous pressure; Class III, pulmonary edema; Class IV, cardiogenic shock. HCT=hematocrit.
PLT=platelet. WBC=white blood cell.
UA=Unstable angina. NSTEMI=non-ST-segment elevation myocardial infarction.
PCI=percutaneous coronary intervention. CABG=coronary artery bypass grafting.
ACEI=angiotensin converting enzyme inhibitors. ARB=angiotensin receptor blocker.

### Clinical outcomes

The 30-day NCB event (all-cause death, reinfarction, nonfatal stroke, or BARC ≥2 bleeding at 30 days) was observed in 136 of the 5430 patients who received fondaparinux (2.5%), in contrast to 110 of the 2636 patients who received LMWH (4.2%) [HR=0.594 (95% CI, 0.462 to 0.764, *P*<0.001] (Fig. 2A and Table 2). The in-hospital NCB event occurred in 91 (3.5%) patients in the LMWH group and in 106 (2.0%) patients in the fondaparinux group [HR=0.581 (95% CI, 0.439 to 0.769), *P*<0.001] (Table 2). At 180 days, the rate of NCB event occurred in 163 (6.2%) patients in the LMWH group and 217 (4.0%) patients in the fondaparinux group [HR=0.637 (95% CI, 0.520 to 0.780), *P*<0.001] (Fig. 2D and Table 2). The rate of MACE (all-cause death, reinfarction, nonfatal stroke) occurred in 72 patients (1.3%) who received fondaparinux and in 45 patients (1.7%) in LMWH at 30 days [HR=0.785 (95% CI, 0.539 to 1.143), *P*=0.207] (Fig. 2B and Table 2). Until the end of the follow-up, we observed that that while the difference failed to reach statistical significance, the fondaparinux group continually showed a trend towards a lower MACE incidence compared to the LMWH group (Fig. 2E and Table 2).

**Figure 1.**
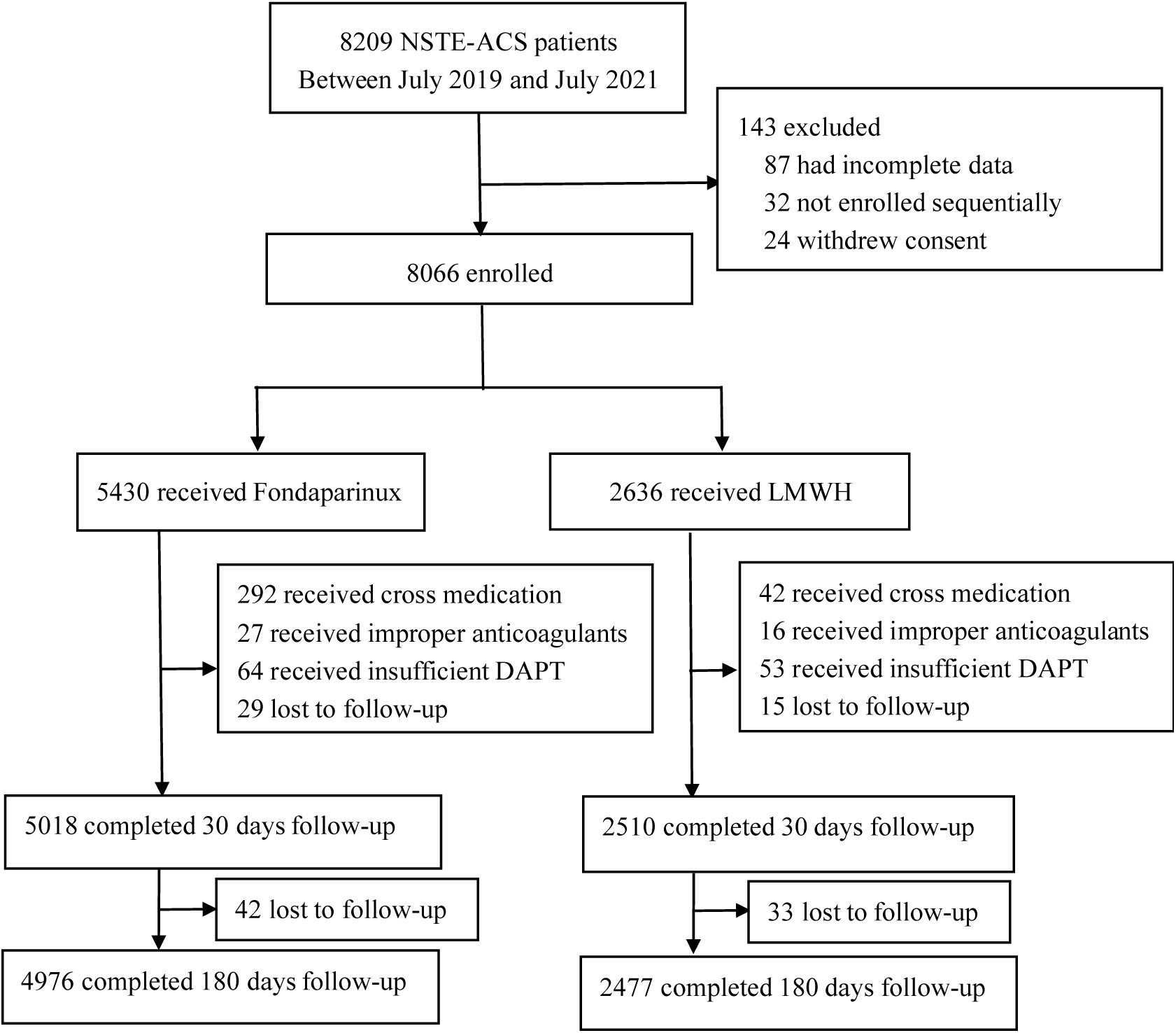
Study flow chart. NSTE-ACS=non-ST-segment elevation acute coronary syndrome. LMWH=low molecular weight heparin. DAPT=dual antiplatelet therapy

**Figure 2.**
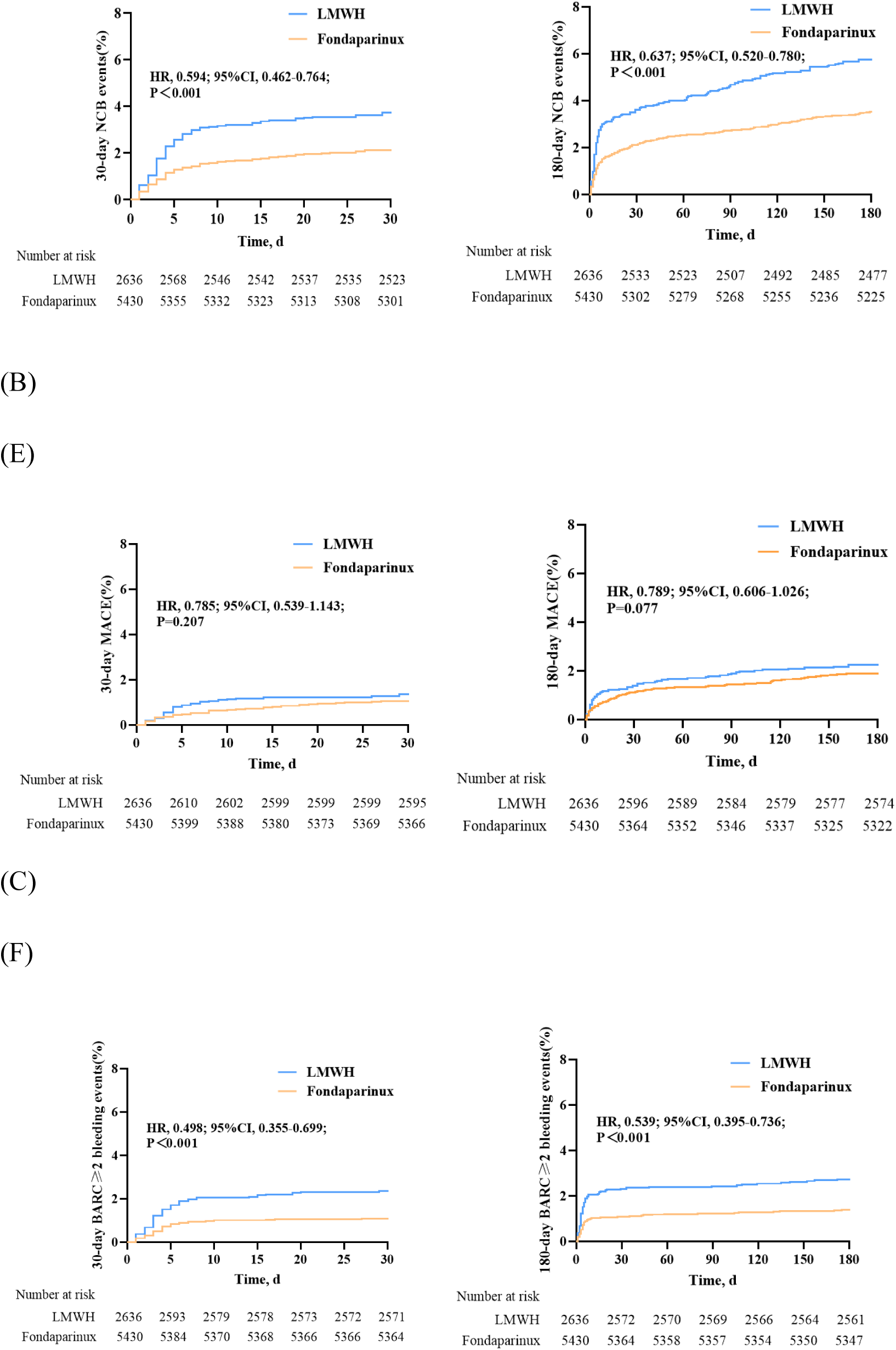
Kaplan-Meier curves for the 30-day and 180-day primary endpoint and its components. (A) 30-Day NCB: The composite of all-cause death, reinfarction, nonfatal stroke, or BARC≥type 2 bleeding. (B) 30-Day MACE: The composite of all-cause death, reinfarction, and nonfatal stroke. (C) 30-Day BARC types 2–5 bleeding. (D) 180-Day NCB: The composite of all-cause death, reinfarction, nonfatal stroke, or BARC≥type 2 bleeding. (E) 180-Day MACE: The composite of all-cause death, reinfarction, and nonfatal stroke. (F) 180-Day BARC types 2–5 bleeding. MACE=major adverse cardiovascular events.

**Table 2:** Clinical outcomes after assignment.

| Events | LMWH<br>(n=2636) | Fondaparinux<br>(n=5430) | HR (95% CI) | p-value |
| --- | --- | --- | --- | --- |
| **In-hospital** |  |  |  |  |
| NCB | 91 (3.5 %) | 106 (2.0 %) | 0.581(0.439-0.769) | < 0.001 |
| MACE | 34 (1.3 %) | 48 (0.9 %) | 0.716(0.461-1.112) | 0.137 |
| all-cause death | 11 (0.4 %) | 17 (0.3 %) | 0.832(0.388-1.782) | 0.636 |
| stroke | 20 (0.8 %) | 29 (0.5 %) | 0.718(0.406-1.270) | 0.255 |
| reinfarction | 3 (0.1 %) | 4 (0.1 %) | 0.677(0.151-3.030) | 0.610 |
| BARC types 2–5<br>bleeding | 58 (2.2 %) | 62 (1.1 %) | 0.533(0.372-0.763) | 0.001 |
| **30 days** |  |  |  |  |
| NCB | 110 (4.2 %) | 136 (2.5 %) | 0.594(0.462-0.764) | < 0.001 |
| MACE | 45 (1.7 %) | 72 (1.3 %) | 0.785(0.539-1.143) | 0.207 |
| all-cause death | 18 (0.7 %) | 30 (0.6 %) | 0.861(0.475-1.561) | 0.622 |
| stroke | 25 (0.9 %) | 34 (0.6 %) | 0.660(0.394-1.106) | 0.115 |
| reinfarction | 3 (0.1 %) | 10 (0.2 %) | 1.550(0.426-5.648) | 0.506 |
| BARC types 2–5<br>bleeding | 66 (2.5 %) | 68 (1.3 %) | 0.498(0.355-0.699) | < 0.001 |
| **180 days** |  |  |  |  |
| NCB | 163 (6.2 %) | 217 (4.0 %) | 0.637(0.520-0.780) | < 0.001 |
| MACE | 91 (3.5 %) | 143 (2.6 %) | 0.789(0.606-1.026) | 0.077 |
| all-cause death | 42 (1.6 %) | 74 (1.4 %) | 0.857(0.587-1.251) | 0.423 |
| stroke | 36 (1.4 %) | 55 (1.0 %) | 0.746(0.490-1.136) | 0.172 |
| reinfarction | 14 (0.5 %) | 23 (0.4 %) | 0.811(0.417-1.577) | 0.537 |
| BARC types 2–5<br>bleeding | 75 (2.8 %) | 84 (1.5 %) | 0.539(0.395-0.736) | < 0.001 |

NCB=all-cause death, stroke, reinfarction, or BARC types 2–5 bleeding
MACE=all-cause death, stroke, reinfarction

The incidence of BARC≥2 bleeding at 30 days was significantly lower in the fondaparinux group compared with the LMWH group [68 cases (1.3%) versus 66 cases (2.5%); HR=0.498, 95% CI, (0.355, 0.699), P<0.001] (Fig. 2C and Table 2).

Similar lower rates of BARC≥2 bleeding were still observed during hospitalization and at six-month follow-up. Specifically, the in-hospital occurrence of BARC≥2 bleeding was substantially lower in the fondaparinux group than in the LMWH group [62 (1.1%) vs. 58 (2.2%), HR=0.533 (95% CI, 0.372 to 0.763), *P*=0.001] (Table 2).

This difference persisted during long-term follow-up, with rates of 84 cases (1.5%) in the fondaparinux group and 75 cases (2.8%) in the LMWH group [HR=0.539 (95% CI, 0.395 to 0.736), *P*<0.001)] (Fig. 2F and Table 2).

### Subgroups

Fondaparinux significantly decreased the risk of ischemia and bleeding in patients with low and intermediate risk scores. For those with a GRACE score ≤140, the Absolute Risk Difference (ARD) for MACE was -0.7% (*P*=0.009), and for bleeding, it was -1.2% (*P*=0.001). For patients with a CRUSADE score ≤40, the ARD for MACE was -0.6% (*P*=0.044), and for bleeding, it was -0.9% (*P*=0.007), thereby enhancing their net clinical benefits. In patients with high bleeding risk profile (CRUSADE score>40), fondaparinux exhibited a significant benefit in reducing bleeding compared to the LMWH group, with an ARD of -3.0% (*P*=0.001) (Fig. 3).

**Figure 3:**
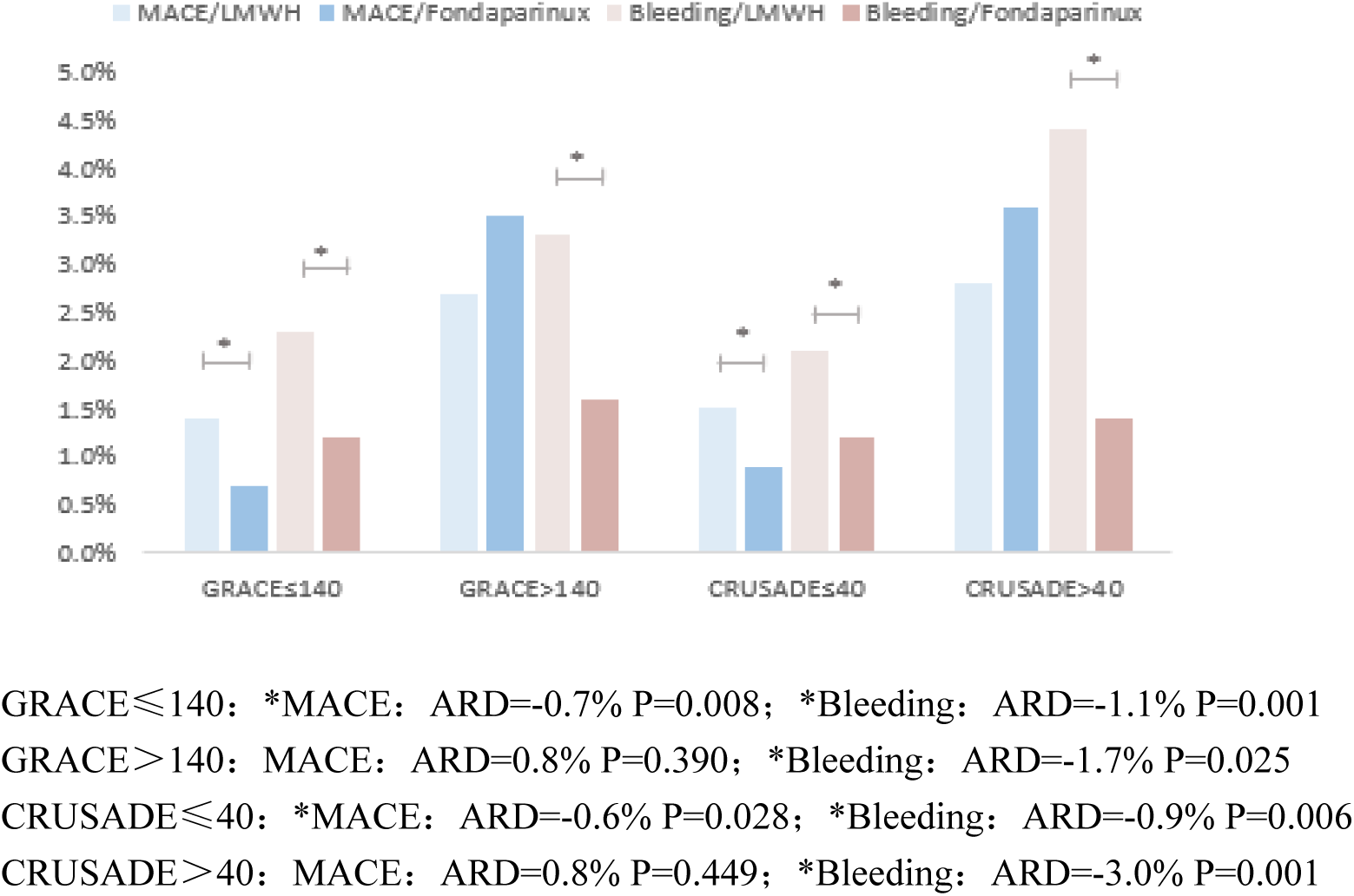
Absolute risk difference (ARD) for LMWH compared with Fondaparinux concerning MACE (all-cause death, stroke, reinfarction) and bleeding (BARC types 2–5) within the four risk stratification. Absolute risk differences (ARDs) are presented: a positive ARD represents the risk increase for Fondaparinux compared to LMWH.

The incidence of the 30-day NCB outcome was consistent across most subgroups. In summary, fondaparinux reduced the incidence of the composite endpoint of bleeding and ischemia events of 30 days compared with LMWH, demonstrating a more positive net clinical benefit (Supplementary Fig. 1).

## Discussion

Our study was a large-scale, real-world clinical trial conducted among Chinese NSTE-ACS patients, exploring the efficacy and safety of fondaparinux relative to LMWH. We also used risk scores to stratify patients to identify the differences in outcomes among patients with different risk stratifications. Our results demonstrated superior efficacy and safety of fondaparinux compared with LMWH. In the group with lower ischemic and bleeding risk, fondaparinux exhibited a greater net benefit compared to the population at higher risk. Among patients at high risk of bleeding, fondaparinux demonstrated enhanced safety performance.

Antithrombotic therapy for patients with NSTE-ACS has evolved significantly over the last 20 years. Initially, there was an emphasis on thrombolytic therapy, which subsequently shifted to combined antithrombotic treatment involving heparin and aspirin[16,17]. The introduction of potent P2Y12 receptor antagonists represents a major advancement[18]. Currently, the standard approach incorporates a combination of dual antiplatelet and anticoagulant agents. The selection and application of parenteral anticoagulants affect the prognosis of patients.

Prior to our investigation, the OASIS-5 trial[7] evaluated the efficacy and safety of fondaparinux relative to enoxaparin, confirming the non-inferiority of fondaparinux. Similarly, the Swedish SWEDEHEART registry study[8] showed reduced bleeding and mortality with fondaparinux in a broader population. The latest guideline[2](Class IB) recommends anticoagulation with fondaparinux in NSTE-ACS patients and suggests UFH to augment anticoagulation during PCI. However, as most of these studies concluded before 2010, their populations and treatments may differ from contemporary cardiovascular practices. The earlier OASIS-5 study[7] did not address the efficacy of dual antiplatelet agents, such as ticagrelor, in patients with ACS, nor did it evaluate the clinical net benefit of triple therapy combining anticoagulants used in ACS with dual antiplatelet agents. In addition, most previous studies focused on European and American populations, with limited research evidence directly from China. Furthermore, risk scores can aid in determining the appropriateness of invasive treatment upon hospital admission and in evaluating ischemia and bleeding risks [2]. There is a lack of attention to how different ischemic and bleeding risk populations might benefit differently from anticoagulant therapy.

To address this data gap, we conducted the present trial, in which a total of 5430 patients received fondaparinux treatment, and 2636 received LMWH treatment. Of these, 4976 and 2477 completed the trial, respectively. Patients received appropriate antiplatelet and other drugs according to clinicians’ discretion. The main findings of our study included the following: (1) fondaparinux treatment led to resulted in a 1.7% absolute reduction (HR 0.594) in the 30-day NCB event rate among patients experiencing MACE or BARC type 2-5 bleeding compared to LMWH; (2) the fondaparinux treatment group demonstrated a 1.2% absolute reduction (HR 0.498) in 30-day BARC ≥ type 2 bleeding compared with the LMWH group; (3) compared with LMWH, the fondaparinux group exhibited lower rates of 30-day all-cause death and stroke, although these differences were not statistically significant. Similarly, the MACE rates were lower in the fondaparinux group but did not reach statistical significance; (4) fondaparinux demonstrated better efficacy and safety in patients at low and intermediate risk compared to those at high risk, and in the high bleeding risk group, it showed greater safety than LMWH.

The PENTUA study[19] evaluated four dosing regimens of fondaparinux (2.5, 4, 8, and 12 mg once daily, subcutaneously) against enoxaparin (1 mg/kg twice daily, subcutaneously) for 3-8 days in ACS patients. The study established 2.5 mg/d as the standard dose for phase 3 studies involving ACS patients. In Accordingly, we adhered to the established dosage, but adjusted the duration of anticoagulation therapy to 2-8 days according to existing guidelines and the OASIS-5 trial [7]. In the OASIS-5 trial, fondaparinux demonstrated non-inferiority to enoxaparin in terms of short term efficacy and significantly reduced bleeding rates. Furthermore, the trial indicated that the decreased, bleeding rate associated with fondaparinux was linked to lower long-term mortality and endpoint event incidence. Analysis of a subgroup of 340 Chinese cases revealed that the combined endpoint and incidence of major bleeding events on the ninth day post-treatment in the fondaparinux group was consistent with global results. Our study showed a lower incidence of ischemic and bleeding events compared to the OASIS-5 trial, which may be related to the higher proportion of patients with unstable angina in our study (69.3% vs. 45.1%). However, compared with the non-inferiority demonstrated by fondaparinux in the OASIS-5 trial relative to enoxaparin, our study indicated that fondaparinux exhibited better efficacy and safety compared to LMWH.

The Swedish SWEDEHEART registry study[8] reached similar conclusions. It found that fondaparinux was associated with a lower risk of bleeding events and death compared with LMWH, both at short-term and long-term follow-up. However, rates of myocardial infarction and stroke were similar. There were similar outcomes in patients with different degrees of renal function and similar results in a subgroup of patients with NSTEMI who had undergone early PCI. In the patients with poor renal function (eGFR ≤ 15mL/min/1.73 m^2^), the efficacy of LMWH was better, but we did not find this result in our research. This discrepancy might be attributable to the relatively better renal function of the patients included in our study.

However, these large studies were conducted and published in the earlier years, and there were relatively fewer studies on the combination of potent P2Y12 receptor antagonists with anticoagulants. The CCC-ACS project[20] found that only one-sixth of patients with NSTE-ACS received care that adhered to high-quality standards, suggesting that evidence-based guidelines were not adequately implemented in the daily clinical practice for Chinese patients with NSTE-ACS.

Therefore, the REFOCAS trial included a broad population of patients with NSTE-ACS and aimed to investigate the outcomes of two anticoagulants in combination with different antiplatelet agents. This approach adhered to existing treatment guidelines and was intended to effectively balance the risks of ischemia and bleeding in these patients, striving for superior clinical outcomes. Notably, the 30-day rates of major adverse cardiovascular and cerebrovascular events, including all-cause death, reinfarction, nonfatal stroke, were lower in both treatment groups in our study compared to those in other multicenter trials[7,8,21,22]. This difference may be attributed to the inclusion of ACS patients with milder syndromes in our study, characterized by a lower mean age, fewer prior infarctions, and a lower prevalence of smoking. Furthermore, the incidence of bleeding events was lower in this study compared with previous large trials[7,21,22]. This may be due to different definitions of major bleeding, as we used BARC ≥ 2 bleeding as the primary safety observation. Moreover, in this study, fondaparinux resulted in fewer bleeding events than LMWH across different follow-up visits. Consequently, it was evident that fondaparinux exhibited a better safety profile than LMWH. However, the differences between studies may not be fully explained by the limited availability of published data from previous large trials. The GRACE risk score[23,24] predicts clinical outcomes and it can be used to estimate the risk of future ischemic events in NSTE-ACS patients, informing decisions regarding the timing of invasive treatments. The CRUSADE score[13,25] offers significant predictive accuracy in assessing risk of major bleeding in ACS patients undergoing coronary angiography. In the OASIS-5 trial, the benefits of fondaparinux were consistent across low-, medium-, and high-risk patients based on the GRACE risk score [26]. However, no study has been found that combines these two risk scores to stratify patients’ ischemia and bleeding risk and to guide in-hospital patients’ anticoagulation therapy. In the low ischemic risk subgroup, the fondaparinux cohort exhibited a decreased incidence of MACE relative to LMWH group. However, this trend was not observed in the high ischemic risk subgroup. Within the low bleeding risk subgroup, the fondaparinux group similarly exhibited a reduction in MACE incidence. Across all four risk subgroups, fondaparinux consistently reduced bleeding risk compared to LMWH, with the most pronounced effect seen in the high bleeding risk subgroup. Our study included approximately 80% non-high-risk ischemic and hemorrhagic patients. In this population, fondaparinux showed superior efficacy and safety than LMWH, likely due to its unique therapeutic mechanism. Therefore, based on these findings, we recommend fondaparinux for patients at a low risk of ischemia and bleeding. In contrast, for patients at a high risk of ischemia, alternative anticoagulants should be considered after a careful balance of ischemia and bleeding risks.

Our study possessed certain limitations. First, the open-label design of the study meant that both clinicians and patients were aware of the patient’s medications, which could introduce potential bias. However, to minimize such biases, an independent CEC adjudicated all endpoint and adverse events. Second, there is currently insufficient theoretical evidence to guide medication decisions based on risk stratification according to GRACE or CRUSADE scores. Large-scale, randomized, controlled clinical trials are necessary to verify the effects of anticoagulant drugs in different risk groups for ischemia and bleeding. Third, all patients have now completed six months of follow-up, and we focused on observing the endpoint event rate at one month. A subsequent one-year follow-up investigation is warranted to determine long-term safety and efficacy.

## Conclusion

In Chinese patients with NSTE-ACS, fondaparinux reduced the incidence of 30- day NCB events of bleeding and ischemia compared with LMWH. Furthermore, fondaparinux demonstrated superior safety in patients with high bleeding risk. Additionally, in the low ischemic and low bleeding risk subgroups, fondaparinux consistently outperformed LMWH in terms of both efficacy and safety.

## Data availability statement

The data underlying this article are available in the article and in its online supplementary material. Further data sharing is available on request from the corresponding author.

